# New insight of adrenal responses in premature neonates versus full term neonates in critical care setting

**DOI:** 10.1101/2020.07.15.20154849

**Authors:** Suzan Abd Razik, Heba Ezzat Hashem, Wafaa Osman Ahmed

**Author notes:** Address correspondence: to Dr. Wafaa osman A Address:-MD, lecturer of Pediatrics’ and neonatology Department - Faculty of Medicine -Ain Shams University, Egypt., /. contributed equally (Suzan Abd Razik, Heba Ezzat Hashem). Declaration of Conflict of Interest: None.

## Abstract

**Background:** Adequate adrenocortical function is essential for survival of critically ill neonates. Although most of them display elevated plasma cortisol concentrations, which reflects activation of the hypothalamic pituitary adrenal axis (HPA), yet; adrenocortical insufficiency is seen in septic shock. Objectives: Evaluate the HPA response in critically ill neonates with shock.

**Methods:** this prospective observational Participant: a total of 60 neonates divided into 3 groups;(A) 30 critical ill neonates with septic shock on inotropic support, (B)15 patients with sepsis with no inotropic support and(C) control group(n=15). Intervention: a single diurnal ACTH reading and two readings for serum cortisol level (diurnal and nocturnal).

**Results:** Gram negative organism was more prevalent among the patients; 53%, 63% in groups A and B respectively. Group A showed Significant statistical hypotension before vasopressor drug administration (p<0.001) as compared to both groups. Group A showed Significant statistical improvement of blood pressure after vasopressor drug administration (p<0.001) as compared to both groups B, C. Serum cortisol was significantly higher in group A(57.21±24.31) and B (48.01±18.27), while it was lower in group C(19.57±16.05). A highly statistically significant rise of serum cortisol level(p=0.000) and ACTH(p=0.000) in group A when was compared to the other two groups.

**Conclusion:** This study introduced a new pattern of serum cortisol response in neonates ranging from very high cortisol level to a near normal values; highlighting a state of glucocorticoid resistance in neonates and relative adrenal insufficiency.

## Introduction

In periods of stress and shock; hypothalamic hormone including corticotropin releasing hormone (CRH) and arginine vasopressin 1 stimulate release of adrenocorticotropin (ACTH) from pituitary; this in turn stimulates release of cortisol from the adrenal glands. Presence of bacterial toxins trigger proinflammatory mediators release that cause neutrophil endothelial cell adhesion together with activation of the clotting mechanisms to form microthrombi. Further arteriolar and arterial vasodilation cause decrease of peripheral vascular resistance and increase cardiac output, leading to warm shock. Later; Blood pressure and cardiac output decrease with or without increase in peripheral vascular resistance, leading to cold shock. It worth mentioning that vasoactive mediators cause distributive defect by shunting of blood and bypassing capillary exchange vessels[1].

Poor capillary perfusion due to shunting and distribution defect together with capillary obstruction by microthrombi cause dysfunction of one or more organs like; kidneys, adrenal glands, brain and heart. In septic shock, inflammatory cytokines including interleukins (IL 1-b and IL-6) and tumor necrosis factor (TNF) may have dual effects on hypothalamic pituitary adrenal axis (HPA); by increasing their hormones initially then, blunting the adrenal axis by the effect of TNF, which contribute to relative adrenal insufficiency(RAI). Relative adrenal insufficiency occur in prolonged states of stress and may result from dysfunction of HPA at any point from hypothalamus to adrenal, and it may contribute to blunted tissue response to glucocorticoids that may appear relatively normal in level. RAI primarily presents with cardiovascular instability in the form of hypotension and shock that may be resistant to fluids and vasopressors [2].

### Subjects and Methods

This study was a prospective descriptive study, that included 60 patients aged below one month (gestational age 32-36weeks), admitted to neonatal intensive care (NICU), Ain Shams university hospital over period of six month from January to June, 2020. Written consents were obtained from care givers of the admitted neonates. The study protocol was approved by Ain Shams University NICU Ethical Committee. Informed consent was taken from mother or legal guardians of included neonates in the study. The sixty neonates were further divided into three groups. Group A: included 30 critically ill patients who are proved clinically and laboratory to have septic shock (15 preterm and 15 fullterm); thus furtherly subdivided into 22(73.3%) patients received double inotropic drug(dopamine and dobutamine on a dose up to 20 mcg /kg/min) and the other 8 patients received both drugs plus nor-adrenaline on a dose of 0.1 to 0.5 mcg /kg/min. Group B: Included 15 patients proved to have sepsis and they did not need inotropic support; (9 preterm, 6 full term)

Group C: 15 health control neonates (8 pretrm, 7 full term), both cases and controls were sex and age matched. Inclusion criteria: hemodynamically unstable neonates. The enrolled babies who developed hypotension and needed inotropic support; the mean arterial pressure < 2 SD for the gestation. Exclusion criteria: suspected cause of adrenal insufficiency, example patients proved to have shock due to: cardiac cause, adrenal cause, ambiguous genitalia, central cause(example; brain malformations, encephalitis, panhypopituitarism), postoperative, postnatal exogenous steroid treatment.

All patients were thoroughly examined after taking full history with emphasis on history of maternal illnesses, gestational age, diagnosis on admission to NICU. The need for respiratory support and inotropes were recorded to facilitate further subgrouping of patients. Evaluation of sepsis markers: Total Leucocytic Count (TLC), neutrophil count, CRP, bacterial blood culture and other cultures. Metabolic profile: Liver function tests, kidney function tests, serum electrolytes, calcium, magnesium and phosphorus, random blood sugar, serum albumin. ACTH and serum cortisol levels: During sepsis total serum cortisol level was measured in neonates with sepsis on inotropic support (double or triple inotropic drugs) twice in the morning (8-9 am) and and in the evening (8-9.30 pm). ACTH was measured once in the morning (8-9 am). Measurement of serum cortisol Using enzyme chemiluminescent technique on the immulite (DPC). Specimen collection through 2mL of venous blood was withdrawn and left to clot. The separated sera were stored at -20°C. The principle of the procedure depends on solid phase competitive chemiluminescent enzyme immunoassay. The solid phase is a polystyrene bead enclosed within an immulite test unit, it is coated with murine anti cortisol antibody. The patient sample and alkaline phosphatase conjugated cortisol are simultaneously introduced into the test unit, and incubated for approximately 30 minutes at 37C with intermittent agitation. During this time, cortisol in the sample competes with the enzyme labeled cortisol for a limited number of antibodies binding sites on the head. Unbound enzyme-conjugate is then removed by a centrifugal wash, after which substrate is added and the test unit is incubated for a further 10 minutes.The chemiluminescent substrate, a phosphate ester of adamantly dioxetane, undergoes hydrolysis in the presence of alkaline phosphatase to yield an unstable intermediate. The continuous production of the intermediate results in the sustained emission of light. The bound complex and thus also the photon output, as measured by the luminometer is inversely proportional to the concentration cortisol in the sample. Reagents:

1. Cortisol test units (100 units): Each barcode-labeled unit contains one head. 1 Kco: 100 units.
2. Cortisol reagent wedge (LCO,) with barcode 6.5 ml alkaline phosphatase (bovine calf intestine) conjugated to cortisol in buffer. Store capped and refrigerated.
3. Cortisol adjustors (LCOL, LCOH): Two vials (low and high), 3mL each, cortisol in processed human serum.

### Measurement of serum adrenocortical hormone (ACTH)

a. Specimen collection:5 cc of venous blood were collected by vein puncture (heamolysis was avoided), the tubes were immersed in an ICP bath. Following collections; plasma was separated from the cells by centrifugation in refrigerated centrifuge, then aliquoted and freezed immediately in plastic tubes at -20C. ACTH test units (LAC1) (100 units). Each barcode labeled unit contains one bead.
b. Principle of the procedure: IMMULITE ACTH is a solid phase, two-site chemiluminescent immunometric assay. The solid phase, a polystyrene bead enclosed within an immulite test unit, is coated with a monoclonal antibody specific for ACTH. While the patient serum sample and alkaline phosphatase-conjugated polyclonal antibody are incubated for approximately 30 minutes at 37C in the test unit with intermittent agitation, ACTH in the sample is bound to form an antibody sandwich complex. Unbound complex is then removed by a centrifugal wash, after which the substrate is added and the Test Unit is incubated for further 10 minutes. The chemiluminescent substrate, a phosphate ester of adamantly dioxetane, undergoes hydrolysis in the presence of alkaline phosphatase to yield an unstable intermediate. The continuous production of these intemediate results in the sustained emission of light, thus improving precision by providing a window for multiple readings. The bound complex and thus also the photon output, as measured by the luminometer is proportional to the concentration of ACTH in the sample. The immulite system automatically handles sample and reagent additions, the incubation and separation step, and measurement of the photon output via the temperature controlled luminometer. It calculates the test results for controls and patient samples from the observed signal, using a stored master curve, and generates a printed report.
c. ACTH reagent wedges (LACA, LACB) LACA: 6.5 mL of an alkaline phosphatase (bovine calf intestine) conjugated to polyclonal rabbit anti-ACTH antibody in buffer. ACTH adjustors (LACL, LACH)Two vials (low and high) of lypophilized ACTH in a bovine protein-based matrix

## Results

### Demographic data

Group A had 15 preterm males and 15 full term females, while group B had 6 full term and 9 preterm babies; 11 of them were females and group C had 7 full term females and 8 preterm males. Three groups were age and sex matched. There was no significant difference between the 3 studied groups regarding mode of delivery, sex and age.

### Laboratory results

Both groups A and B had significantly high total leukocytic count, neutrophil and CRP as compared with the control group. Reverse correlation between neutrophil count and cortisol level was documented. Both ACTH and cortisol are not affected by addition of inotropes in group A.

### Culture proved sepsis

Sepsis proved cultures was evident in 27 out of 45 patients of both groups A(19) and B(8), where Gram positive staph aureus and E coli had the highest proportion of culture yield (25% each), followed by both streptococci and pneumococci (7.1% each) and finally was the klebsiella proved cultures in (3.6%) of patients.

### Blood pressure in the 3 groups

Systolic blood pressure was nearly equal in group B and C (mean=70.2± 6.44), (71.93±9.6) respectively, while it was lower in group A (46.6±6.58). So as the diastolic pressure; that was nearly in the same range; (41.6±7.3), (41.93±5.39) in groups B and C respectively, while group A had lower diastolic range (23.39±3.88). Although blood pressure was improved after addition of inotropes (group A); yet both diastolic and systolic were lower than other 2 groups.

### Cortisol level assessment in the 3 groups

Serum cortisol (both readings) was higher in both groups A, B as compared with group C. While ACTH was nearly in the same range in both groups B and C; yet it was found to be higher (double time) in group A rendering a significant statistical difference. A characteristic pattern of cortisol release was documented in group A; 73.33% had quantitative folding (am>pm values with concomitant rise), 50% had quantitative folding and disturbed rhythm (am>pm values with inconcomitant rise), 23.33% had quantitative folding with reversed rhythm (pm>am) and 10% had quantitative low level (am was within normal range, pm slightly increased)

## Discussion

### culture proved sepsis

Group A had culture proved sepsis in 19(63.3%); of which 42% was gram negative, while group B had culture proved sepsis in 8(53%); of which all were gram negative. E coli was the most prevalent followed by the klebsiella species. This is supported by Macharashivii et al. [3] and Couto et al. [4] who reported 18-78% of neonatal sepsis were due to gram negative organism.

### Blood pressure difference among the three groups

Non statistical difference in blood pressure was found comparing group B and C. While hypotension was evident in group A before and after inotropic support in comparison to the other two groups showing a highly statistical significant difference, indicating a state of a peripheral tissue resistance expressing a vasopressor resistant hypotension that was more pronounced in group A especially preterm patients. Table 1

**Table(1):**
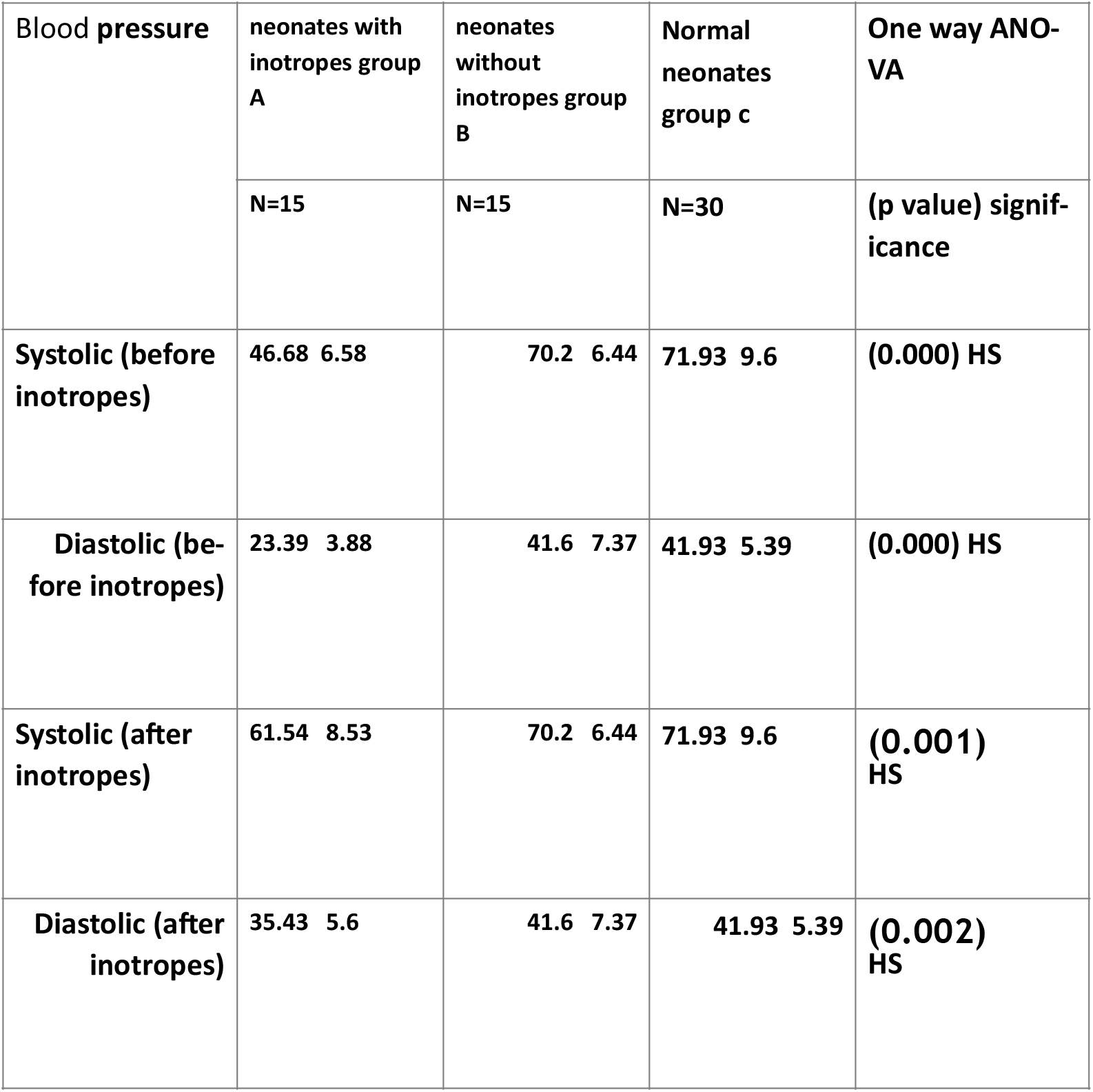
Comparison between the studied groups regarding blood pressure parameters before and after inotropic support This table shows a highly statistically significant decrease in mean value of the systolic and diastolic blood pressure for group A as compared to the other two groups (B,C) even after inotropic support.

| Blood pressure | neonates with inotropes group A | neonates without inotropes group B | Normal neonates group c | One way ANOVA |
| --- | --- | --- | --- | --- |
|  | N=15 | N=15 | N=30 | (p value) significance |
| Systolic (before inotropes) | 46.68 6.58 | 70.2 6.44 | 71.93 9.6 | (0.000) HS |
| Diastolic (before inotropes) | 23.39 3.88 | 41.6 7.37 | 41.93 5.39 | (0.000) HS |
| Systolic (after inotropes) | 61.54 8.53 | 70.2 6.44 | 71.93 9.6 | (0.001) HS |
| Diastolic (after inotropes) | 35.43 5.6 | 41.6 7.37 | 41.93 5.39 | (0.002) HS |

To demonstrate the response of premature babies versus full term ones regarding vasopressor drugs (groupA); a total number of 30 ill neonates was equally divided into preterm and full term, where 11 of each group received double inotropic support while the other 4 received triple inotropic support. The difference of response of full term and preterm neonate, in face of vasopressor drugs, can be shown by improved systolic blood pressure in full term post inotropic support as compared to preterm of same group(p<0.05). So full term babies showed a statistical significant rise in systolic blood pressure compared to preterm patients.

In addition, after starting inotropic support, we found preterm patients of group A had statistically significant hypotension as compared to those in group B, while on the other hand a non statistically significant difference was observed among full term patients of same groups(A,B), indicating a more poor response to vasopressors and immature hemodynamic compensatory mechanisms in pre terms. This goes on with Biniwale et al. [5] who described a vasopressor resistance hypotension in pre terms. Table 2

**Table(2):**
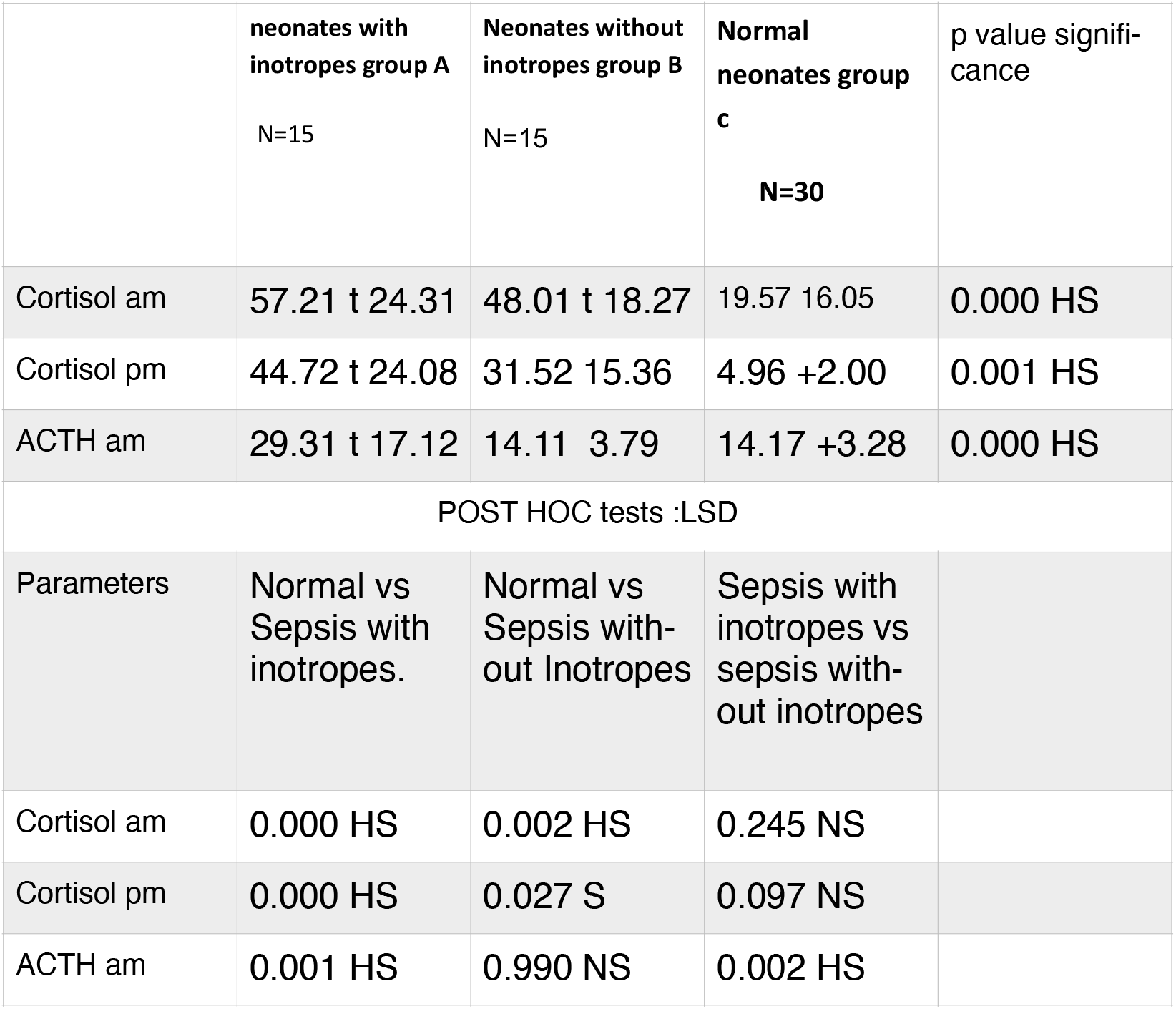
Comparison between all the studied groups regarding serum cortisol level and ACTH. Normal level of ACTH=10-60 pg/ml. Normal serum cortisol level =5-25 ug/dl This table shows a highly statistically significantdecrease in mean value of the systolic and diastolic blood pressure for group A compared to the other two groups (B, C) even after inotropic support.

This confirmed the poor response to vasopressor and an immature hemodynamic compensatory mechanisms, also Higgins et al. [6] stated that some preterms are either vasopressor resistant and/ or dependant due to the relative adrenal insufficiency plus, down regulation of cardiovascular adrenergic receptors due to both critical illness and chronic stimulation. As one of the genomic actions of corticosteroids is the upregulation of those mentioned receptors and the down regulation thought to be due to RAI

Although absolute adrenal insufficiency is quite rare in critically ill neonates, relative adrenal insufficiency is relatively common. Together with the presence of hemodynamic instability despite adequate fluids and vasopressors; other diagnostic clues include hyponatremia, hyperkalemia and hypoglycemia [7].

In the present study, on monitoring the laboratory clues for adrenal insufficiency, hyperkalemia was statistically significant in group A as compared to the other two groups, while hyponatrmia and hypoglycaemia showed a non statistical difference. This agrees with one study [8] who reported a hyperkalemic state in ill preterm presenting with inadequate adrenal insufficiency. But it is important to take into consideration that electrolyte assessment was not taken in serious evaluation because the ill groups (A,B) were receiving continuous fluid therapy in NICU. So in this study, laboratory evidence of adrenal insufficiency was assessed basically on measuring total serum cortisol level.

### Assessing serum cortisol level in the 3 groups

In assessing the total serum cortisol level in all the studied groups, the following data were recorded; First, normal neonates (group C) showed a circadian rhythm of an (am to pm) ratio (3:1) in both preterms and full terms, with a mean cortisol level (19.57ug/ dl) and a range of (3.5-35ug/dl) in am values and range of (2.9-6.9ug/dl) in pm values. Table 3. In addition, there was non signficant difference in serum cortisol and ACTH levels comparing normal preterm and normal full terms. Opposite to this, Vermes et al. [9], mentioned that diurnal rhythms were still absent in first 2 months, and the typical circadian rhythms of the plasma cortisol levels were present in infants aged 3 months. Second, total serum cortisol level shows a highly statistical significant rise in both group A and B together with loss of the recorded rhythm in group C (normal neonates). In agreement with that two studies [10,11]; confirmed the normal circadian rhythm of cortisol secretion, (characterized by achievement of the highest concentration in the morning and subsequently decreased nadir in the evening), is lost during critical illness. This also goes on with study done by Asare [12] who stated that for critically ill patients, it is not necessary to obtain cortisol levels at a specific time of the day because most patients lose the diurnal variation in their cortisol levels.

In the current study it was also found that in group A (septic neonates with inotropic support), a quantitative folding of mean total serum cortisol level together with dysrythmic pattern were detected, categorizing this group into four response patterns. Category I: 15/30(50%) quantitative folding shows of both (am and pm) values with a concomitant rise. Category II: 5/30(16.6%) quantitative and reversed rhythm shows quantitative folds of both (am and pm) values but with a pm values higher than am values. Category III: 3/30(10%) quantitative and disturbed rhythm shows quantitative folds than normal of am values while pm values showed a relatively lower folding values compared to the previous two categories. Finally, Category IV: 7/30(23%) (relative low) where am values fall within normal range values despite severe sepsis, i.e; relative low serum cortisol. In this category preterm represents (71%) and full term 28.5%.

Passing through the adrenal gland response in the four categories gives an idea about the state of the hypothalamic pituitary axis (HPA) activation behaviour in sepsis and septic shock, where a stage of initial rise occurs in serum cortisol level exemplified in category (I&II), followed by exhaustion of the adrenal gland, exemplified in category (III&IV). One study8 reported that patients with severe sepsis have cortisol values three fold higher than normal and that very preterm neonates would be anticipated to be at increased risk for cortisol insufficiency in the face of acute illness and stress.

Relative adrenal insufficiency is present when the cortisol response is inadequate for the patient’s degree of stress, despite a cortisol level that may appear normal. This is due to dysfunction at any point in the HPA axis or due to resistance to glucocorticoids [2]. Asare et al [12] defined Relative adrenal insufficiency as a random cortisol level of less than 25 ug/dl. That random cortisol testing is a useful diagnostic threshold for the diagnosis of adrenal insufficiency.

In the present study, aiming at assessing the severity of the vasopressor resistant shock based on adrenal gland response (serum cortisol level), it was found that triple inotropic therapy was used in (4/8) 50% in category I (highest folding), (2/8) 25% in category IV (relatively low) followed by 12.5% in other two categories (III&IV). Very high levels in category I also indicates a state peripheral tissue resistance to glucocorticoids, while the relatively low category (IV) indicates a state of an adrenal exhaustion. This goes on with the study done by Annane et al. [13] who mentioned that, the hypothalamus-pituitary-adrenal axis is activated, initially leading to increased free cortisol. Next, the entire axis is exhausted and peripheral cortisol resistance leads to the development of adrenal insufficiency.

This also agrees with [14], who described a state of adrenal exhaustion in chronic critical illness due to catecholamine receptor desensitization or down regulation and/or chronic secretion of cytokines and other substances that suppress the HPA axis.

In the present study, it was concluded that relative adrenal insufficiency can be either in a quantitative form as presented in (relative low) category IV (mean serum cortisol 19 ug/dl) “adrenal exhaustion”, or as a functional adrenal insufficiency despite high cortisol levels indicating “peripheral tissue cortisol resistance”. Conforming the state of ‘tissue resistance”, mean serum cortisol level in preterms (55.13ug/dl) and full terms (59.29 g/dl) in group A (septic neonates with inotropic support) showed a significant and highly significant reversed correlation respectively to both systolic and diastolic blood pressure. This goes on with one study 2, who found that relative adrenal insufficiency is present when the cortisol response is for the patient’s degree of stress, despite a cortisol level that ever that may appear normal and was explained by dysfunction at any point in the HPA axis or due to resistance to glucocorticoids. Studies [15,16] also reported that an inadequate HPA axis response to stress can aggravated by peripheral resistance to glucocorticoids. In addition to reduction of glucocorticoid receptors and an increase in the conversion of cortisol to inactive cortisone by enhanced activity of 11-B hydroxysteroid dehydrogenase stimulated by IL-2, IL-4, and IL-13. Many studies [17,18,4] believed that a random cortisol level in severely stressed patients (ie. with hypotension) should be above 25 mcg/dl and that higher levels may be appropriate in patients with septic shock due to “tissue cortisol resistance. Studies [8,18] recorded that random cortisol value of <15 mcg/dl is considered to be a clear evidence of adrenal insufficiency. Also Soliman et al. [19] reported that 11 of 30 (36.6%) septic term new-borns had basal cortisol values of <15 mcg per 100 ml.

Dellinger et al. [20] reported that relative adrenal insufficiency is seen in 16.3 to 55% of the patients with septic shock. While other studies [13,21,22] reported that prevalence rates of adrenal insufficiency (AI) in critically ill patients vary according to the characteristics of the study population and the diagnostic criteria used and rates as high as 60-90% have been documented in patients with septic shock. One study [23] reported that the proportion of relative adrenal insufficiency in children with sepsis (66.6%) was much higher than that without sepsis (25%), which declares the strong association between sepsis and septic shock with relative adrenal insufficiency.

### Cortisol versus blood pressure in both preterm and full term

There was reversed significant correlation between serum cortisol level and both systolic and diastolic blood pressure in preterm and fullterm of group A. This finding could demonstrate the intact HPA axis while reflects the resistance at the tissue levels. While we reported a significant correlation between ACTH and serum cortisol in full term (p=0.00 7); the same correlation was not found in preterm group(p=0.68).

Studying serum ACTH behaviour in all the studied groups, it was concluded that ACTH showed a highly statistically significant rise in group A, (septic neonates with inotropic support) compared to the other two groups (group B septic neonates not receiving inotropes and group C normal neonates). Table 3. The high mean ACTH (29.3pg/dl) together with the high mean cortisol level (57.2ug/dl) in group A (septic neonates with inotropic support) (a positive correlation) indicates either a direct stimulation of the HP (hypothalamic pituitary axis) provoked by the septic inflammatory mediators in case of severe sepsis and septic shock (a hypothalamic pituitary dependent mechanism), loss of the normal negative feedback mechanism (negative feedback block) between the adrenal and pituitary glands, indicating a central resistance of the hypothalamic/pituitary to the adrenal glucocorticoids. A poor adrenal response to ACTH can be also a contributing factor. These factors collectively lead to the creation of a pseudo positive feedback loop between pituitary and adrenals.

The non significant levels of ACTH in group B (septic neonates without inotropic support) reaching (14.17 pg/dl) nearly the normal values in group C (14.1 pg/dl), despite the rise in serum cortisol level (mean 48ug/dl), indicates that the initial rise m serum cortisol in response to sepsis may occur as “a pituitary independent mechanism” in cases not reaching the septic shock state, or may be in part due to reduction in negative feedback effect of cortisol on hypothalamic pituitary axis (negative feedback block).

This goes on with Sharaga et al [22], who mentioned that although severe stress activates the HPA axis, dissociation may occur between plasma ACTH and cortisol concentrations, as demonstrated by suppressed ACTH and elevated cortisol concentrations. Addingly, a study done by Boonen et al. [24], suggested a pituitary independent mechanism for increased cortisol production during critical illness. In group A, number of inotropes used, sex and mode of delivery showed non statistical significant correlation with serum cortisol level and ACTH. In group A; studying the drugs that may be aggravating the state of relative adrenal insufficiency: antiepileptic drugs (phenytoin) and antifungal (fluconazole) usage was reported. 5/30(16.6%) received antifungal therapy (fluconazole) and 8/30(26.6%) received anti epileptic (phenytoin), 2 out of these 8 were preterm and showed a relatively low serum cortisol level (category IV). Kawai and Ichikawa. [25] mentioned that, phenytoin, barbiturates, and rifampicin are drugs that accelerate the metabolism of cortisol and most synthetic glucocorticoids by inducing hepatic mixed-function oxygenase enzymes, causing adrenal insufficiency in patients with limited pituitary or adrenal reserve and in agreement with this Nicolas et al. [26], also, mentioned an association between high-dose fluconazole and adrenal insufficiency in critically ill patients.

Both serum cortisol and ACTH level were not affected by mode of delivery, sex or the inotropic support that was added in group A.

## Conclusion

Septic shock causes a state of relative adrenal insufficiency that contributes to the higher mortality rate. Early steroid measurement and replacement can improve vassopressor resistance and hypotension leading to a better clinical outcome.

## Data Availability

the data are available

What is already known?

Adrenal insufficiency in critically ill neonate

What is added by the study?

Different cortisol secretion pattern in the neonates either preterm or fullterm

## Reference

[1] Vrezas I, Willenberg HS, Bornstein SR (www.Endotext) Chapter 1, Adrenal Cortex. Development Anatomy, Physiology USMLE Step 1 Physiology Lecture Notes; 2013; pp. 263–289

[2] Gomez-Sanchez CE Adrenal dysfunction in critically ill patients. N Engl J Med; 2013; 368:1547–1549. (doi: 10.1056/NEJMe1302305).

[3] Macharashvili N, Kourbatova E, Butsashvili M, et al. Etiology of neonatal blood stream infections in Tbilisi, Republic of Georgia. Int J Infect Dis;2009; 13(4):499–505. (doi: 10.1016/j.ijid.2008.08.020).

[4] Renato C Couto 1, Elaine A A Carvalho, Tânia M G Pedrosae et al., A 10-year prospective surveillance of nosocomial infections in neonatal intensive care units. Am J Infect Control; 2007; 35(3):183–9. doi: 10.1016/j.ajic.2006.06.013.

[5] Biniwale M, Sardesai S and Seri I Steroids andVasopressor-Resistant Hypotension in Preterm Infants. Current Pediatric Reviews, 2013; 9(1): 75–83.

[6] Higgins S, Friedlich P, Seri I Hydrocortisone for hypotension and vasopressor dependence in preterm neonates: a meta-analysis. Journal of Perinatology, 2010; 30(6):373–378. (doi: 10.1038/jp.2009.126)

[7] Sprung CL, Annane D, Keh D, et al. Corticus Study Group.Group.Hydrocortisone therapy for patients with septic shock. NEngl J Med; 2008; 358:111–124.(doi: 10.1056/NEJMoa071366).

[8] Fernandez EF and Watterberg KL Relative adrenal insufficiency in the preterm and term infant. Division of Neonatology, Department of Pediatrics, University of New Mexico School of Medicine, Albuquerque, NM, USA Journal of Perinatology; official Journal of the California Perinatal; 2009;29:S44–S49. (doi: 10.1038/jp.2009.24).

[9] Vermes I and Beishuizen A The hypothalamic-pituitary-adrenal response to critical illness. Best Pract Res Clin Endocrinol Metab; 2001;15:495–511. (doi: 10.1053/beem.2001.0166)

[10] B Venkatesh 1, R H Mortimer, B Couchman, Evaluation of random plasma cortisol and the low dose corticotropin test as indicators of adrenal secretory capacity in critically ill patients: a prospective study. Anaesth Intensive Care. 2005 Apr;33(2):201–9. doi: 10.1177/0310057X0503300208.

[11] J Nijm, M Kristenson, A G Olsson, L Jonasson. Impaired cortisol response to acute stressors in patients with coronary disease. Implications for inflammatory activity. J Intern Med. 2007 Sep;262(3):375–84. DOI: 10.1111/j.1365-2796.2007.01817.x

[12] Asare K Diagnosis and Treatment Adrenal PatientInsufficiency in the Critically ill patient. Pharmacotherapy; 2007;27(11):1512–1528. (doi: 10.1592/phco.27.11.1512)

[13] Annane D, Maxime V, Ibrahim F, et al., Diagnosis of adrenal insufficiency in severe sepsis and septic shock. Am J Respir Crit Care Med; 2006;174:1319–1326. (doi: 10.1164/rccm.200509-1369)

[14] Arnildo LJ, Eduardo AR, Luiza KM, et al. Adrenal insufficiency in children with sepsis. Pediatric Intensive Care Unit; 2011;23(4):478–483. (doi.org/10.1590/S0103-507X2011000400013).

[15] Pringent NG, C Lee, C Lam, et al. Transient adrenocortical insufficiency of prematurity and systemic hypotension in very low birthweight infants. Arch Dis Child Fetal Neonatal Ed. Mar; 2004; 89(2): F119–F126. doi: 10.1136/adc.2002.021972

[16] Indyk JA, Candido-Vitto C, Wolf IM, et al. Reduced glucocorticoid receptor protein expression in children with critical illness. Horm ResPaediatr; 2013; 79: 169–178. (doi: 10.1159/000348290. Epub 2013 Mar 8).

[17] Mark Endocrine Development - Adrenal Glands UNSW. 2011. Embryology- www.embryology.med.Unsw.edu.com

[18] Cooper MS and Stewart PM Adrenal insufficiency in critical illness. J Inten sive Care Med; 2007; 22(6):348–62.(doi.org/10.1177/0885066607307832).

[19] Soliman AT, Taman KH, Rizk MM, et al., Circulaing adrenocorticotropic hormone (ACTH) and cortisol concentrations in normal, appropriate for gestational age newborns versus those with sepsis and respiratory distress: cortisol response to low-dose and stan-dard-dose ACTH tests. Metabolism; 2004; 53:209–214.

[20] R. Phillip Dellinger, Mitchell M. Levy, Jean M. Carlet,, et al. 2008 Surviving Sepsis Campaign: International guidelines for management of severe sepsis and septic shock: 2008. Intensive Care Med 34, 17–60. (doi.org/10.1007/s00134-007-0934-2)

[21] Hebbar KB, Petrillo T, Fortenberry JD 2012 Adrenal insufficiency and response to corticosteroids in hypotensive critically ill children with cancer. J Crit Care. Epub Jun 12; 2012;27:480–487. (doi: 10.1016/j.jcrc.2012.03.013).).

[22] Shraga LY 2013 Critical illness-related corticosteroidinsufficiency in children. Hormone Research in Paediatrics, 80(5): 309–317.

[23] Elfaramawy A. Hepatoadrenal syndrome in Egyptian children with liver cirrhosis with and without sepsis. EJMHG 2012;13: 337–42.

[24] Boonen E, Vervenne H, Meersseman P et al., Reduced cortisol metabolism during critical illness. N Engl J. 2013; 368:1477-1488.L.(doi: 10.1056/NEJMoa1214969. Epub 2013 Mar 19.).

[25] S Kawai 1 and Y Ichikawa Drug interactions between glucocorticoids and other drugs, 1994 Mar;52(3):773–8.

[26] Nicolas CN, and Evangelia C, George PC Adrenal Insufficiency. 2011 ch13 endotext, the most accessed source on endocrinology for medical professionals.

